## Supplementary Table 1-3 for "Engineered phage endolysin eliminates *Gardnerella* in bacterial vaginosis without damaging the healthy vaginal microbiome"

**Supplementary Table 1:** Various concentrations of PM-477 and incubation times were tested on vaginal smear samples to establish the protocol (see Methods). *Gardnerella* and all bacteria were detected by FISH co-staining and the mean *Gardnerella* cell number covering 10 epithelial cells was determined.

| **Treatment** | **Baseline** | **Puffer** | | | **2 µg/ml PM-477** | | | **20 µg/ml PM-477** | | | **200 µg/ml PM-477** | | |
|---|---|---|---|---|---|---|---|---|---|---|---|---|---|
| **Incubation time** | 0h | 2h | 6h | 24h | 2h | 6h | 24h | 2 h | 6h | 24h | 2h | 6h | 24h |
| **No. of *Gardnerella* cells** | 4 200 | 4 000 | 3 900 | 3200 | 4300 | 4200 | 4200 | 4200 | 3500 | 2300 | 4400 | 2000 | 150 |

**Supplementary Table 2:** Wild-type endolysin genes identified in prophage-like regions of *Gardnerella* genomes. The identification number, the annotation, the predicted protein sequence and the total amino acid length are presented for each gene.

| Name | NCBI Accesion No. | Protein ID | Protein sequence | Total length (aa) |
|---|---|---|---|---|
| **EL1** | NC_017456<br>CP002725.1 | AEF31373.1 | MSKKGIDVSEWQGDIDFNAVKASGVEFVIIRAGYGIGCKDKWFE<br>QNYRKAKTAGLDVGAYWYSYANSGFEAAEEAQSLMNMLSGKSFE<br>YPVYFDLEEKSQLNRGRAFCDSLITSFCNKLEACGYYAGFYTSL<br>STANNLVSAHVRNRYALWIAQWNTHCNYQGSYGLWQYSSNGSVP<br>GVAGRVDMDYAYVDYPSIIKNAGLNGCKNGGSDQAARTSSIDEV<br>AREVINGAWGNGSTRKQRLTSAGYDYASVQNKVNELLGVKACRK<br>SVDELAREVIRGAWGNGSTRKQRLAQAGYDYDTVQKRVNELL | 306 |
| **EL2** | NZ_LWSP010<br>00031.1<br>Contig_31 | WP_065189413.1 | MSKRGIDVSEWQGDIDFNAVKASGVEFVIIRAGYGIGCKDKWFE<br>QNYRKAKTCGLDVGAYWYSYANSGFEAAEEAQSCVNMLSGKSFE<br>YPVYFDLEEKSQLNRGRAFCDSLITGFCNKLESCGYYAGFYTSL<br>STANNLVSAHVRNRYALWIAQWNTHCSYQGSYGLWQYSSSGSVP<br>GVAGRVDMDYAYVDYPSIIKNVGLNGCKNGGSDQAARTSSIDEV<br>AREVINGAWGNGNERKQRLTSAGYDYASVQNKVNELLGVKACRK<br>SVDEIAREVIRGTWGNGSTRKQRLTQAGYDYDTVQKRVNELL | 306 |
| **EL3** | NZ_LWSP010<br>00130.1<br>Contig_130 | WP_065189629.1 | MSKKGIDVSVWQGDIDFNSVKASGVEFVIIRAGYGIGHKDKWFE<br>ENYRKAKTAGLDVGSYWYSYASSAGEASEEAQSCVNILSGKSFE<br>YPIYFDLEEKSQLNRGRDFCDSLITSFCNKLEACGYYAGFYTSL<br>SVANNLVSSHVRDRYALWIAQWNTHCSYQGSYGLWQYSSSGSVN<br>GIAGRVDMDYAYVDYPSVIKNAGLNGYQNGGSYTAPQTSSIDEV<br>AREVINGDWGNGNDRKNRLISAGYDYASVQNKVNELLGVKAYRK<br>SVDELAREVIRGTWGNGSMRKHRLTQAGYDYDAVQKRVNELL | 306 |
| **EL4** | NZ_ADAN01<br>000092.1<br>Contig00100 | WP_004111234.1 | MSKKGIDVSEWQGDIDFNAVKASGVEFVIIRAGYGIGCKDKWFE<br>QNYRKAKTCGLDVGAYWYSYANSGFEAAEEAQSCVNMLSGKSFE<br>YPVYFDLEEKSQLNRGRAFCDSLITSFCSKLETYGYYAGFYTSL<br>SVVNNLVSAHVRDRYALWIAQWNTHCSYQGSYGLWQYSSSGSVP<br>GVAGRVDMDYAYVDYPSIIKNVGLNGCKNGGSDQATRTSSIDEV<br>AREVINGAWGNGNERKQRLTSAGYDYASVQNKVNKLLGVKAYRK<br>SVDELAREVIRGTWGNGNERKQRLAQAGYDYDTVQKRVNELL | 306 |
| **EL5** | ATJR0100016<br>6.1<br>Cont321.1 | EPI54247.1 | MSKKGIDVSEWQGDIDFNAVKASGVEFVIIRAGYGIGCKDKWFE<br>QNYRKAKTAGLDVGAYWYSYANSSSEAAEEAQSCANMLSGKSFE<br>YPVYFDLEEKSQLNRGRAFCDSLITGFCSKLEACGYYAGFYTSL<br>STANNLVSAHVRNRYALWIAQWNTHCSYQGSYGLWQYSSNGSVP<br>GVAGRVDMDYAYKDYPSIIKNAGLNGCKNGGSDQAARTSSIDEV<br>AREVINGAWGNGNERKQRLTQAGYDYTSVQNKVNKLLGVKACRK<br>SVDELAREVIRGTWGNGNERKNRLTQAGYDYDTVQKRVNELL | 306 |

| | | | | |
|---|---|---|---|---|
| **EL6** | ADER010000<br>28.1<br>Contig_00038 | EIK79883.1 | MSKKGIDVSVWQGDIDFNAVKASGVEFVIIRAGYGIGCKDKWFE<br>QNYRKAKTCGLDVGAYWYSYANSGFEAAEEAQSCVNMLSGKSFE<br>YPVYFDLEEKSQLNRGRAFCDSLITSFCSKLESCGYYAGFYTSL<br>STANNLVSAHVRNRYALWIAQWNTHCDYQGSYGLWQYSSSGSVP<br>GVAGRVDMDYAYKNYPSIIKNAGLNGCKNGGSNQAARTSSIDDV<br>AREVINGAWGNGNERKQRLTQAGYDYASVAK | 251 |
| **EL7** | NZ_ADAM01<br>000002.1<br>Contig001 | WP_004104952.1 | MSKKGIDVSEWQGDIDFNAVKASGVEFVIIRAGYGIGCKDKWFE<br>QNYRKAKTAGLDVGAYWYSYANSASEAAEEAQSCANMLSGKSFE<br>YPVYFDLEEKSQLNRGRAFCDSLITSFCSKLETYGYYAGFYTSL<br>STANNLVSSHVRNRYALWIAQWNTHCSYQGSYGLWQYSSSGSVP<br>GVAGRVDMDYAYKDYPSIIKNAGLNGCKNGGSDQAARTSSIDEV<br>AREVINGAWGNGNERKQRLTQAGYDYTSVQNKVNKLLGVKACRK<br>SVDELAREVIRGTWGNGNERKNRLTQAGYDYDTVQKRVNELL | 306 |
| **EL8** | ATJO0100000<br>3.1<br>cont2.1 | EPI52092.1 | MSKKGIDVSEWQGDIDFNAVKASGVEFVIIRAGYGIGCKDKWFE<br>QNYRKAKTAGLDVGAYWYSYANSSSEAAEEAQSCVNMLSGKSFE<br>YPVYFDLEEKSQLNRGRAFCDSLITSFCSKLETYGYYAGFYTSL<br>STANNLVSSHVRNRYALWIAQWNTHCSYQGSYGLWQYSSNGSVP<br>GVAGRVDMDYAYVDYPSIIKNAGLNGYKNGGSYTAPQTSSIDDV<br>AREVINGAWGNGNERKQRLTQAGYDYTSVQNKVNKLLGVKACRK<br>SVDELAREVIRGTWGNGNERKNRLTQAGYDYDTVQKRVNELL | 306 |
| **EL9** | NZ_ADEN010<br>00017.1<br>ctg00024 | WP_004118369.1 | MSKKGIDVSEWQGDIDFNAVKASGVEFVIIRAGYGIGCKDKWFE<br>QNYRKAKTCGLDVGAYWYSYANSGFEAAEEAQSCVNMLSGKSFE<br>YPVYFDLEEKSQLNRGRVFCDSLITSFCNKLEACGYYAGFYTSL<br>STANNLVSSHVRNRYALWIAQWNTHCDYQGSYGLWQYSSSGSVP<br>GVAGRVDMDYAYVDYPSIIKNAGLNGCKNGGSDQAARTSSIDEV<br>AREVINGAWGNGSTRKQRLTSAGYDYASVAK | 251 |
| **EL10** | NZ_LXJL0100<br>0047<br>Contig_47 | WP_064340486.1 | MSKRGIDVSVWQGDIDFNAVKASGVEFVIIRAGYGIGHKDKWFE<br>ENYRKAKTVGLDVGAYWYSYASSAGEASEEAQSCVNILSGKSFE<br>YPVYFDLEEKSQLNRGRDFCDSLITSFCNKLEACGYYAGFYTSL<br>SVANNLVSSHVRDRYALWIAQWNTHCSYQGSYGLWQYSSSGSVN<br>GIAGRVDMDYAYVDYPSVIKNAGLNGYKNGGSYTAPQISSIDEV<br>AREVINGDWGNGNERKQRLTSAGYDYASVQNKVNELLGVKAYRK<br>SVDELAREVIRGTWGNGSTRKQRLTQAGYDYNAVQKRVNELL | 306 |
| **EL11** | ATJN0100006<br>1.1<br>Cont71.1 | EPI51560.1 | MSKRGIDVSVWQGDIDFNAVKASGVEFVIIRAGYGIGHKDKWFE<br>QNYRKAKTTGLDVGAYWYSYASSAGEAAEEAQSCVNILSGKSFE<br>YPVYFDLEEKSQLNRGRDFCDSLITSFCNKLETYGYYAGFYTSL<br>SVANNLVSSHVRDRYALWIAQWNTHCDYQGSYGLWQYSSSGSVD<br>GIAGRVDMDYTYVDYPSVIKKAGLNGYKNGGSYTAPQTSSIDEV<br>AREVINGDWGNGNERKNRLTSAGYDYTSVQNKVNELLGVKAYRK<br>SVDELAREVIRGTWGNGSTRKQRLTQAGYDYDAVQKRVNELL | 306 |
| **EL12** | KMT47143.1<br>contig00001 | WP_048730021.1 | MSKKGIDVSVWQGDIDFNAVKASGVEFVIIRAGYGIGHKDKWFE<br>ENYRKAKTAGLDVGSYWYSYASSAGEVALEAQSCVNILSGKSFE<br>YPVYFDLEEKSQLNRGRDFCDSLITSFCNKLEACGYYAGFYTSL<br>SVANNLVSSHVRDRYALWIAQWNTHCSYQGSYGLWQYSSSGSVN<br>GIAGRVDMDYAYVDYPSVIKNAGLNGYQNGGSYTAPQTSSIDEV<br>AREVINGDWGNGIERKNRLTSAGYDYTSVQNKVNELLGVKAYRK<br>SVDELAREVIRGTWGNGKTRKQRLTQAGYDYNAVQKRVNELL | 306 |
| **EL13** | APW18536.1 | WP_076002731.1 | MSKKGIDVSVWQGDIDFNAVKASGVEFVIIRAGYGIECKDKWFE<br>QNYRKAKTAGLDVGAYWYSYANSGFEAAEEAQSCVNMLSGKSFE<br>YPVYFDLEEKSQLNRGRAFCDSLITSFCNKLESCGYYAGFYTSL<br>STANNLVPAHVRNRYALWIAQWNTHCDYQGSYGLWQYSSSGSVP<br>GVAGRVDMDYAYVDYPSIIKNAGLNGYKNGESHQATRTTSIDEV<br>AREVINGAWGNGNERKQRLTQAGYDYASVQNKVNELLGVKACRK<br>SVDELAREVIRGTWGNGNERKNRLTSAGYDYDTVQKRVNELL | 306 |
| **EL14** | APW18794.1 | WP_076002856.1 | MSKKGIDVSVWQGDIDFNAVKASGVEFVIIRAGYGIGCKDKWFE<br>QNYRKAKTCGLDVGAYWYSYANSGFEAAEEAQSCVNMLSGKSFE<br>YPVYFDLEEKSQLNRGRAFCDSLITSFCNKLEACGYYAGFYTSL<br>STANNLVSAHVRNRYALWIAQWNTHCSYQGSYGLWQYSSSGSVP<br>GVAGRVDMDYAYVDYPSIIKNAGLNGYKNGESHQATRTTSIDEV<br>AREVINGAWGNGNERKQRLTSAGYDYASVQNKVNELLGVKACRK<br>SVDELAREVIRGAWGNGSTRKQRLTSAGYDYDTVQKRVNELL | 306 |

**Supplementary Table 3:** Bacterial cell density before and after treatment with PM-477 as well as buffer control, as determined by FISH microscopy. The number of bacterial cells per ten exfoliated epithelial cells in the vaginal swabs of all 15 BV patient samples are presented. The method of calculation is explained in the M&M section.

| Patient samples | ***Gardnerella*** | | | | ***L. iners*** | | | | ***L. crispatus*** | | | | ***A. vaginae*** | | | |
|---|---|---|---|---|---|---|---|---|---|---|---|---|---|---|---|---|
| | **Baseline** | **Control** | **PM-477** | **Octenisept** | **Baseline** | **Control** | **PM-477** | **Octenisept** | **Baseline** | **Control** | **PM-477** | **Octenisept** | **Baseline** | **Control** | **PM-477** | **Octenisept** |
| 1 | 4200 | 3200 | 150 | | 150 | 80 | 20 | | 0 | 0 | 0 | | 0 | 0 | 0 | |
| 2 | 500 | 500 | 300 | | 0 | 0 | 0 | | 0 | 0 | 0 | | 0 | 0 | 0 | |
| 3 | 5600 | 5000 | 3000 | | 0 | 0 | 0 | | 1 | 0 | 0 | | 0 | 0 | 0 | |
| 4 | 1500 | 1000 | 100 | | 200 | 200 | 20 | | 0 | 0 | 0 | | 50 | 20 | 500 | |
| 5 | 1000 | 300 | 300 | | 0 | 0 | 0 | | 0 | 0 | 0 | | 200 | 0 | 500 | |
| 6 | 2500 | 3000 | 200 | | 500 | 800 | 400 | | 20 | 100 | 100 | | 200 | 200 | 200 | |
| 7 | 9000 | 7500 | 4000 | | 60 | 60 | 20 | | 0 | 0 | 0 | | 1000 | 1000 | 500 | |
| 8 | 1100 | 900 | 600 | | 0 | 0 | 0 | | 0 | 0 | 10 | | 50 | 0 | 100 | |
| 9 | 800 | 800 | 1 | | 5 | 0 | 0 | | 0 | 0 | 0 | | 3 | 0 | 1 | |
| 10 | 500 | 300 | 500 | | 0 | 0 | 0 | | 0 | 0 | 0 | | 0 | 0 | 0 | |
| 11 | 5200 | 5200 | 30 | 100 | 150 | 100 | 20 | 0 | 0 | 0 | 0 | 0 | 0 | 0 | 0 | 0 |
| 12 | 1200 | 600 | 300 | 100 | 0 | 0 | 0 | 0 | 0 | 0 | 0 | 0 | 100 | 30 | 30 | 3 |
| 13 | 4500 | 2000 | 600 | 1500 | 0 | 0 | 0 | 0 | 0 | 0 | 0 | 0 | 400 | 400 | 600 | 0 |
| 14 | 650 | 300 | 1 | 0.5 | 60 | 50 | 50 | 0 | 0 | 0 | 0 | 0 | 40 | 20 | 40 | 0 |
| 15 | 600 | 800 | 1 | 10 | 120 | 60 | 20 | 4 | 0 | 0 | 0 | 0 | 100 | 120 | 80 | 0 |
| Median | **1200** | **900** | **300** | **100** | **5** | **0** | **0** | **0** | **0** | **0** | **0** | **0** | **50** | **0** | **40** | **0** |
